# Novel integrase mutations linked to genotypic DTG resistance in non-B HIV-1 strains from African participants: The DTG RESIST study

**DOI:** 10.1101/2025.10.23.25338481

**Authors:** N Han, T Loosli, M Sauermann, İ Çelikağ, N Anderegg, B C Baye, C Bolton Moore, L Buzaalirwa, H Byakwaga, C Chimbetete, P V Ebasone, S Goodrich, J Huwa, C Kasozi, A Mafoua, A C Massamba, E Messou, A Minga, G Murenzi, G Muula, W Muyindike, S J Naidoo, D M Nsonde, A Poda, R Ramdé, A Semeere, L Singh, H F Günthard, M Egger, J Giandhari, R Lessells, R D Kouyos, the DTG RESIST Study Group

## Abstract

**Background:** Integrase mutations associated with dolutegravir resistance have been well characterized, but based on limited data from non-B subtypes.

**Objectives:** We aim to identify integrase mutations not currently classified as integrase strand transfer inhibitor (INSTI) resistance mutations (DRMs) in individuals with viremia on dolutegravir-based regimens.

**Methods:** Mutations in integrase sequences in the DTG RESIST study from African countries were detected using Stanford HIVdb v9.8. We used a viral genome-wide association study (GWAS) approach to identify mutations not classified as major or accessory INSTI DRMs but associated with dolutegravir resistance. We performed the same GWAS on drug-naïve sequences from the Los Alamos HIV-1 database to identify mutations associated with viraemia under DTG exposure.

**Results:** Among 387 sequences, 107 (27.6%) showed at least intermediate dolutegravir resistance. Fourteen integrase mutations not classified as major or accessory DRMs (S39R, L45I, I72L, L74I, V79I, F100Y, I113V, S119R, V126A, K156N, Q177L, I208M, A265V, and R284G) were significantly associated with resistance. V79I (adjusted odds ratio [aOR] 169.2, 95% credible interval [CrI] 18.2–2871.4) and I72L (aOR 67.7, 95% CrI 7.1–1326.2) were strongly associated with resistance. S39R, L45I, I72L, L74I, V79I, F100Y, S119R, and K156N were linked to established INSTI resistance pathways, and I72L, L74I, V79I, V126A, and K156N were associated with viraemia under DTG exposure.

**Conclusions:** We identified several integrase mutations outside established DRM categories that are strongly associated with dolutegravir resistance. Dolutegravir resistance evolution is complex; likely involves mutations not currently classified as DRMs.

## Introduction

Dolutegravir (DTG), a second-generation integrase strand transfer inhibitor (INSTI), has been shown to be non-inferior to efavirenz-based regimens in terms of viral suppression, and to have advantages in safety, tolerability, and ease of use.^1–3^ Furthermore, DTG’s high barrier to resistance may help preserve treatment options in patients who experience antiretroviral therapy (ART) failure.^4^ Consequently, the WHO recommended DTG in 2018 as the preferred antiretroviral agent for most people living with HIV (PWH).^5^ Since then, global scale-up efforts have led to its widespread adoption in national treatment guidelines as the preferred first-line and second-line treatment for all PWH.^6^

Despite DTG’s high genetic barrier to resistance, recent cohort studies and national surveys have reported emerging resistance worldwide.^7–9^ Although the prevalence of drug resistance among PWH with viraemia varied across settings, resistance is typically associated with well-characterized drug resistance mutations (DRMs) in the HIV-1 integrase gene, such as R263K, G118R, N155H, and Q148H/R/K,^10^ which are classified as major INSTI DRMs by the Stanford HIVdb algorithm^11^ and by the IAS-USA.^12^ Phenotypically, R263K and N155H cause a modest, about 2-fold reduction in DTG susceptibility, G118R a strong, 19-fold reduction, whereas Q148H/R/K has only minimal effect unless combined with other resistance mutations.^10^

Previous studies have identified some accessory or minor integrase mutations, such as L74I, M50I, and K156N, that may also influence viral replication capacity and contribute to resistance^13,14^ yet their role in the absence of major mutations seems rather limited.^15^ It is unclear whether these mutations reduce DTG susceptibility on their own, act synergistically with major DRMs, or contribute to the evolution of high-level resistance by enhancing replication fitness. Identifying other mutations is therefore crucial for understanding the evolutionary pathways of HIV-1 and its capacity to adapt under drug pressure from INSTIs.

In this study, we identified integrase mutations associated with DTG resistance that are not currently classified as major or accessory DRMs in the Stanford HIVdb algorithm.^11^ We analysed data from the DTG RESIST study,^16^ a large international cross-sectional study investigating DTG resistance in individuals experiencing virological failure on DTG-based ART. By characterizing these mutations in a diverse clinical population, all of whom were infected with non-B subtypes, we aim to provide new insights into the evolutionary mechanisms underlying DTG resistance.

## Methods

### Ethics

Ethics approval was received by the University of Bern from the Cantonal Ethics Committee of Bern (Ref. No: 2021-01504); by the University of Kwazulu-Natal, South Africa (Ref No: BREC/00005146/2022), and all participating sites.

### Study setting

The DTG RESIST study is a large multicentre cross-sectional study nested within the International epidemiology Databases to Evaluate AIDS (IeDEA).^17^ From June 2022 to May 2025, it enrolled adults and adolescents living with HIV in Africa, Asia, and Latin America who experienced virological failure while receiving DTG-based ART.^16^ Participants were eligible if they had been on DTG for at least three months and had at least one routine viral load measurement above 1000 copies/mL. All participants provided informed consent. Study teams collected demographic and clinical data, and whole blood samples for plasma and/or dried blood spot (DBS) preparation. The African samples were shipped to the KwaZulu-Natal Research Innovation and Sequencing Platform in Durban, South Africa.^18^ We performed sequencing if the viral load on the enrolment sample was above 1000 copies/mL.

### Sequencing and participant characteristics

Sanger sequencing was performed on the plasma or DBS samples using the HIV-1 Genotyping kit with Integrase (ThermoFisher Scientific Inc., Waltham, MA, USA), and sequences were analysed using the Stanford HIVdb algorithm (version V9.8).^11^ We categorized mutations in the integrase region as major, accessory, or other mutations based on the Stanford HIVdb algorithm. We used Rega (version 3.47)^19^ and COMET (version 2.4)^20^ to assign HIV-1 subtypes, prioritizing classification based on the integrase region. If Rega failed to assign a subtype from integrase, subtyping was based on the protease and reverse transcriptase regions using Rega; if no subtype could be assigned with Rega, we used the COMET classification for the integrase region. Only sequences from Africa were included because, at the time of analysis (September 2025), sequences from the other regions were not yet available. If the participant was enrolled more than once in the study, only the sequence from the first enrollment was included for the analysis.

We compared sequence and participant characteristics with and without DTG resistance using Welch’s two-sample t-test to compare discrete variables (number of major, accessory, and other mutations) and chi-squared test for categorical variables (HIV-1 subtypes and country with study sites). DTG resistance was defined as at least intermediate level of DTG resistance scores predicted by the Stanford HIVdb algorithm.

### Genome-wide association study (GWAS)

We performed a genome-wide association study (GWAS) to identify other integrase mutations associated with DTG resistance.^21^ After sequence alignment, a genetic distance matrix was constructed and analysed by principal coordinates analysis (PCoA).^22^ We included the top ten principal components (PCs) as covariates in the GWAS model to account for population structure. GWAS was performed using Firth logistic regression,^23^ restricted to mutations observed more than three times. We controlled multiple testing using the Benjamini-Hochberg (BH) procedure, with statistical significance defined as an adjusted p-value of < 0.05.^24^

An additional GWAS identified other integrase mutations associated with viraemia under DTG exposure. We used sequences from the DTG RESIST, comprising individuals who experienced virological failure under DTG-based treatment, and drug-naïve sequences from the Los Alamos HIV-1 database, retrieved on 25 August 2025.^25^ To match the subtype distribution, we excluded subtype B sequences from the Los Alamos dataset as sequences from the DTG RESIST did not contain subtype B. Pairwise genetic distances were calculated to construct a distance matrix between all sequences. We then further selected three genetically similar drug-naïve control sequences for each viraemic DTG RESIST sequences applying the Hungarian algorithm.^26^ This is to minimize the total genetic distance between DTG RESIST and Los Alamos sequences, while controlling for inter-dataset variability. As in the previous GWAS conducted to identify other mutations associated with resistance, we considered only mutations observed at least three times in the dataset. We applied Firth logistic regression, adjusting for the population structure, and used the same BH correction and significance threshold (adjusted p-value < 0.05) to identify significant mutations for viraemia under DTG exposure.

### Post-GWAS exploratory analyses

To further quantify the impact of the GWAS-identified other integrase mutations on DTG resistance, we applied two complementary approaches: Fisher’s exact test and the Boruta feature-selection algorithm.^27^ Fisher’s exact test was used to identify other integrase mutations overrepresented in sequences with DTG resistance compared to those without, with statistical significance defined as a Bonferroni-adjusted p < 0.05. The Boruta algorithm, based on a random forest model trained with both original and permuted (i.e., randomly shuffled) variables, was used to identify mutations that were predictive of DTG resistance, defined as those with importance scores exceeding the maximum among permuted variables.^27^ We also examined the co-occurrence of GWAS-identified integrase mutations in sequences with DTG resistance using the pairwise Phi coefficient.

We employed a ridge-penalized Bayesian logistic regression models to assess the direction of the associations (positive vs. negative) between the identified mutations and DTG resistance, using three models: (i) unadjusted, (ii) adjusted model with the first ten PCs from the GWAS model that identified mutations associated with resistance, and (iii) adjusted models with the PCs and demographical and clinical variables including HIV-1 subtype, country of origin, sex, number of accessory INSTI DRMs, time on DTG-based treatment, and documented exposure to first-generation INSTIs. Their associations were also examined when sequences were stratified by HIV-1 subtypes. We further assessed the distribution of the identified mutations across signature DTG mutations which comprise mutational pathways using Fisher’s exact test. These pathways represent distinct evolutionary trajectories by which HIV-1 acquire mutations in the integrase gene that reduce DTG susceptibility. We compared four mutually exclusive mutational pathways at four amino acid positions including G118R, Q148HRK, N155H, and R263K.^10^

## Results

### Study population

We included a total of 387 HIV-1 sequences from individuals experiencing viraemia while receiving DTG-based ART. Subtype C (n = 220, 56.8%) was the most prevalent subtype, followed by subtypes G or CRF 02_AG (n = 68, 17.6%), and A1 (n = 56, 14.5%). 107 sequences (27.6%) had predicted intermediate or high-level resistance to DTG. Sequences with DTG resistance had a median of 3 (range: 1-5) major and 1 (range: 0-3) accessory INSTI DRM. The average number of other integrase mutations was higher in sequences with DTG resistance (20, range: 12-28) compared to those without (18, range: 9-31) (Table 1, Supplementary Figure S1).

**Table 1.**
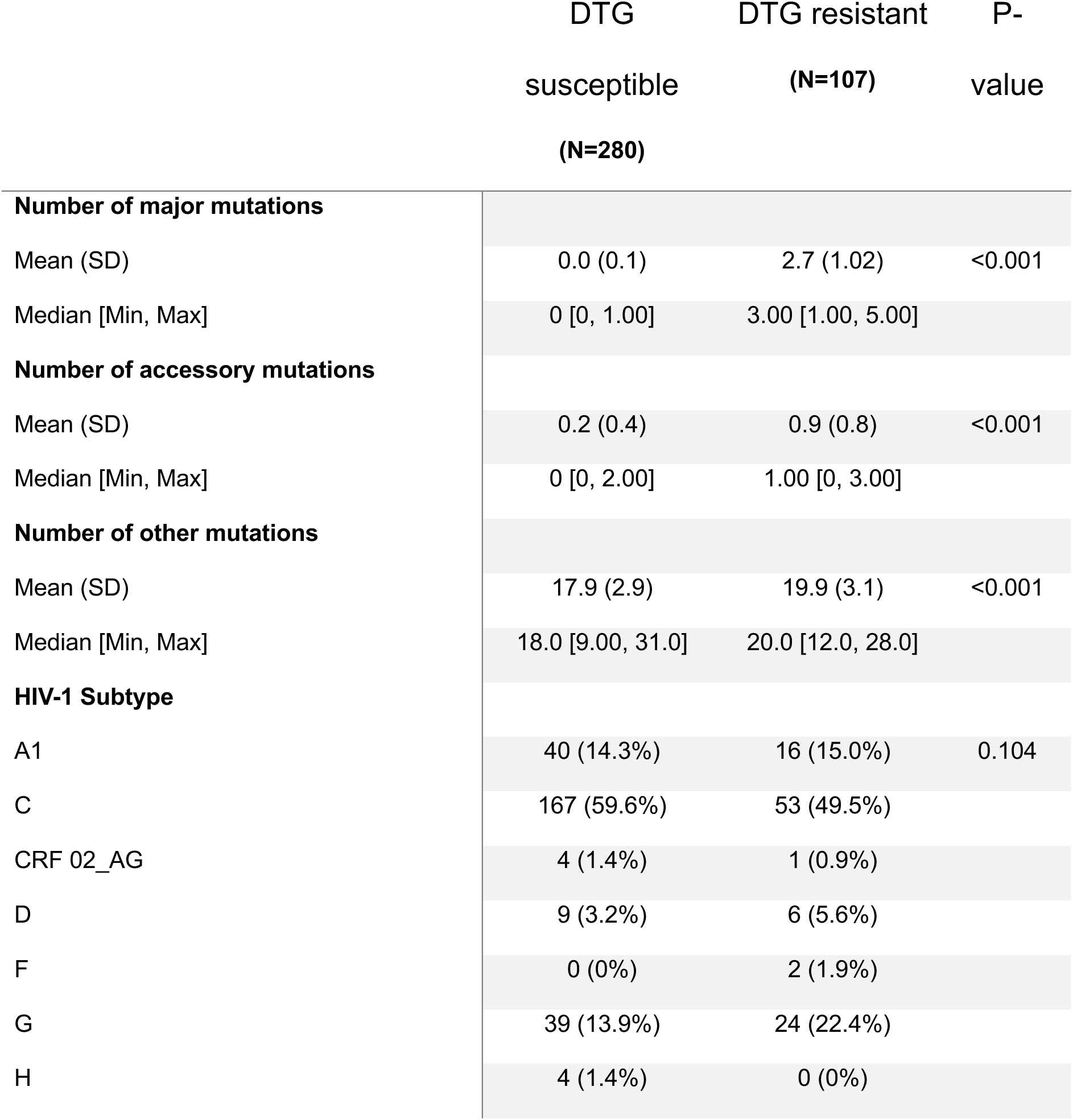
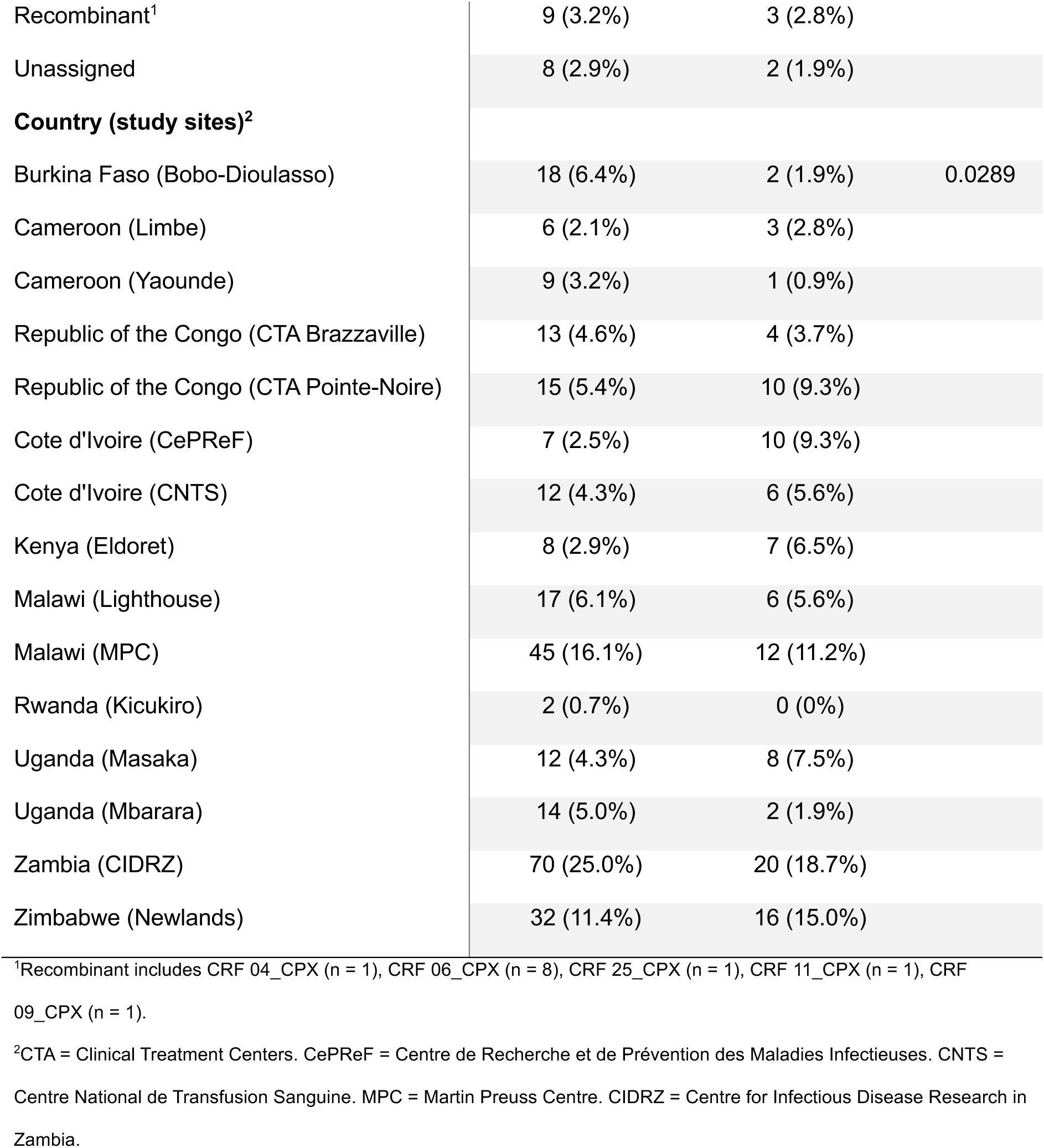
Description of sequences with DTG resistance and without. DTG resistance was defined as sequences with at least one intermediate level of major INSTI DRM. P-value was calculated using a two-sample t-test for numerical variables (number of major, accessory, other mutations) and a chi-squared t-test for categorical variables (HIV-1 subtypes, country with study sites). DTG = dolutegravir. INSTI = integrase strand transfer inhibitor. DRM = drug resistance mutation.

|  | DTG<br>susceptible<br>(N=280) | DTG resistant<br>(N=107) | P-value |
| --- | --- | --- | --- |
| **Number of major mutations** |  |  |  |
| Mean (SD) | 0.0 (0.1) | 2.7 (1.02) | <0.001 |
| Median [Min, Max] | 0 [0, 1.00] | 3.00 [1.00, 5.00] |  |
| **Number of accessory mutations** |  |  |  |
| Mean (SD) | 0.2 (0.4) | 0.9 (0.8) | <0.001 |
| Median [Min, Max] | 0 [0, 2.00] | 1.00 [0, 3.00] |  |
| **Number of other mutations** |  |  |  |
| Mean (SD) | 17.9 (2.9) | 19.9 (3.1) | <0.001 |
| Median [Min, Max] | 18.0 [9.00, 31.0] | 20.0 [12.0, 28.0] |  |
| **HIV-1 Subtype** |  |  |  |
| A1 | 40 (14.3%) | 16 (15.0%) | 0.104 |
| C | 167 (59.6%) | 53 (49.5%) |  |
| CRF 02_AG | 4 (1.4%) | 1 (0.9%) |  |
| D | 9 (3.2%) | 6 (5.6%) |  |
| F | 0 (0%) | 2 (1.9%) |  |
| G | 39 (13.9%) | 24 (22.4%) |  |
| H | 4 (1.4%) | 0 (0%) |  |
| Recombinant<sup>1</sup> | 9 (3.2%) | 3 (2.8%) |  |
| Unassigned | 8 (2.9%) | 2 (1.9%) |  |
| **Country (study sites)<sup>2</sup>** |  |  |  |
| Burkina Faso (Bobo-Dioulasso) | 18 (6.4%) | 2 (1.9%) | 0.0289 |
| Cameroon (Limbe) | 6 (2.1%) | 3 (2.8%) |  |
| Cameroon (Yaounde) | 9 (3.2%) | 1 (0.9%) |  |
| Republic of the Congo (CTA Brazzaville) | 13 (4.6%) | 4 (3.7%) |  |
| Republic of the Congo (CTA Pointe-Noire) | 15 (5.4%) | 10 (9.3%) |  |
| Cote d'Ivoire (CePReF) | 7 (2.5%) | 10 (9.3%) |  |
| Cote d'Ivoire (CNTS) | 12 (4.3%) | 6 (5.6%) |  |
| Kenya (Eldoret) | 8 (2.9%) | 7 (6.5%) |  |
| Malawi (Lighthouse) | 17 (6.1%) | 6 (5.6%) |  |
| Malawi (MPC) | 45 (16.1%) | 12 (11.2%) |  |
| Rwanda (Kicukiro) | 2 (0.7%) | 0 (0%) |  |
| Uganda (Masaka) | 12 (4.3%) | 8 (7.5%) |  |
| Uganda (Mbarara) | 14 (5.0%) | 2 (1.9%) |  |
| Zambia (CIDRZ) | 70 (25.0%) | 20 (18.7%) |  |
| Zimbabwe (Newlands) | 32 (11.4%) | 16 (15.0%) |  |

<sup>1</sup>Recombinant includes CRF 04\_CPX (n = 1), CRF 06\_CPX (n = 8), CRF 25\_CPX (n = 1), CRF 11\_CPX (n = 1), CRF 09\_CPX (n = 1).
<sup>2</sup>CTA = Clinical Treatment Centers. CePReF = Centre de Recherche et de Prévention des Maladies Infectieuses. CNTS = Centre National de Transfusion Sanguine. MPC = Martin Preuss Centre. CIDRZ = Centre for Infectious Disease Research in Zambia.

### Association of other integrase mutations with DTG resistance

We identified 14 other integrase mutations significantly associated with genotypic DTG resistance using the GWAS approach, including S39R, L45I, I72L, L74I, V79I, F100Y, I113V, S119R, V126A, K156N, Q177L, I208M, A265V, and R284G (Figure 1). When comparing prevalence of other integrase mutations in sequences with Fisher’s exact test, 25 mutations including eight (S39R, I72L, V79I, V126A, K156N, Q177L, I208M, and A265V) of the 14 GWAS-identified mutations were significantly overrepresented among sequences with DTG resistance compared to those without (Supplementary Table 1). For four (S39R, I72L, V79I, and Q177L) of the eight overrepresented GWAS-identified mutations, odds ratios were infinite because they were absent in sequences without resistance. Of the 14 mutations identified in the GWAS, ten (S39R, L45I, I72L, V79I, S119R, V126A, K156N, Q177L, I208M, and A265V) were also included in the list of mutations considered predictive of DTG resistance by the Boruta feature-selection algorithm (Supplementary Figure 2). Finally, two mutation pairs (I72L-K156N and V79I-R284G) from the 14 GWAS-identified mutations showed significant pairwise associations (Supplementary Figure 3). Both associations were weakly positive with Phi coefficients of 0.27 and 0.28, respectively.

**Figure 1.**
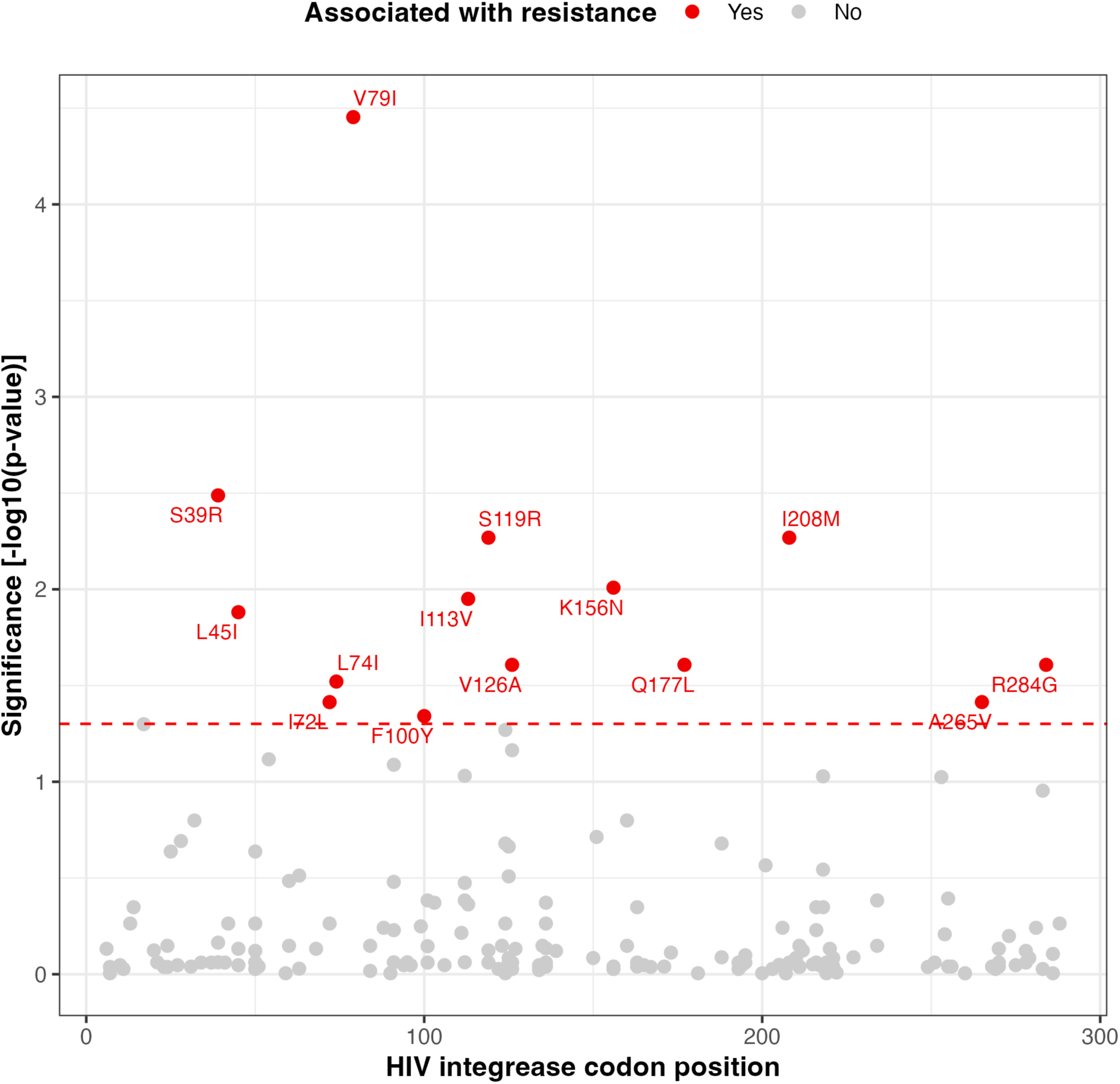
Genome-wide association study (GWAS) of other integrase mutations and associations with genotypic DTG resistance. For GWAS, Firth logistic regression was applied, including the first 10 PCs and subtypes to account for population structure. P-values were adjusted using the BH procedure, with a significance threshold of adjusted p-value < 0.05. The name of significant mutations were displayed and mutations identified in the GWAS for association with DTG resistance (Figure 1) are highlighted in red. DTG = dolutegravir. PC = principal component. BH = Benjamini-Hochberg.

### Adjusted analyses and mutational pathways

In the Bayesian logistic regression models adjusting for population structure (PCs from the GWAS) and demographic and clinical variables, seven (I72L, L74I, V79I, V126A, K156N, I208M, and R284G) of the 14 mutations were significantly positively associated with DTG resistance, whereas one (A265V) showed a significant negative association (Figure 2). The V79I mutation showed the strongest positive association with an adjusted odds ratio (aOR) of 169.2 (95% credible intervals (Crl) = 18.2-2871.4), followed by I72L (aOR = 67.7, 95% Crl = 7.1-1326.2).

**Figure 2.**
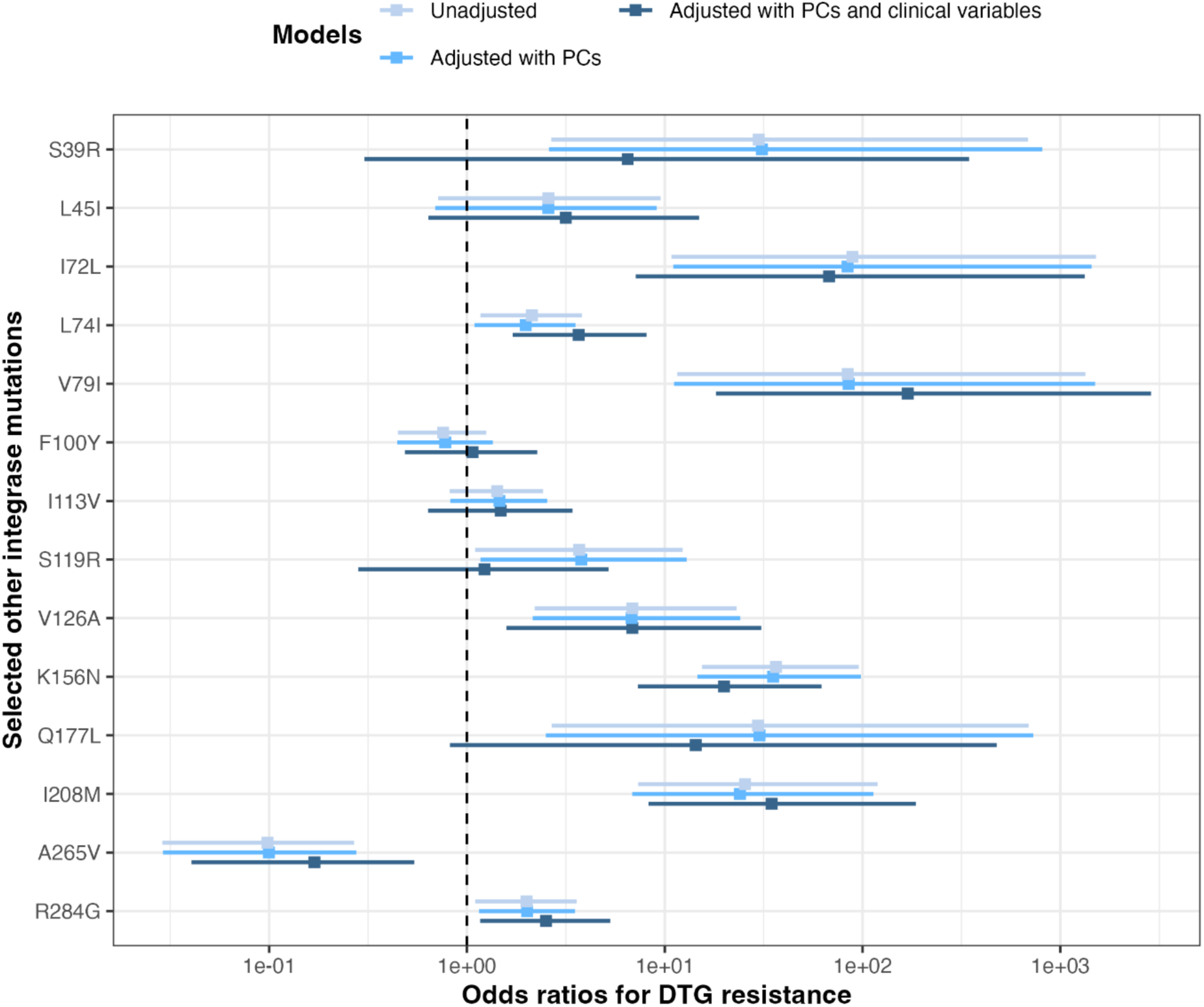
Odds ratios for DTG resistance. Odds ratios with 95% credible intervals were calculated using a Bayesian regression model. Mutations identified from GWAS firth regression were included in the analysis. Different models are in different colours. Unadjusted model included each other integrase mutation without adjusting for any other variables. Different covariates were added into different models: (i) adjusted model with PCs include the first ten PCs used in the GWAS firth regression to adjust for population structure, (ii) adjusted model with PCs and clinical variables include the first ten PCs and clinical variables including HIV-1 subtype, country, sex, number of accessory INSTI DRMs, time on DTG-based treatment, and documented exposure to first-generation INSTI. DTG = dolutegravir. GWAS = genome-wide association study. PC = principal component. INSTI = integrase strand transfer inhibitor. DRMs = drug resistance mutations.

Most associations between mutations and DTG resistance were consistent across HIV-1 subtypes (Supplementary Figure 4). However, comparisons were limited due to small sample sizes in subtypes A1 and G or AG. Eight of the 14 mutations (S39R, L45I, I72L, L74I, V79I, F100Y, S119R, and K156N) were associated with different mutational pathways (Figure 3). Particularly, for the V79I mutation, all DTG resistant cases (n = 12) occurred with the Q148HRK pathway. Similarly, for the L45I mutation, all five resistant cases occurred with R263K.

**Figure 3.**
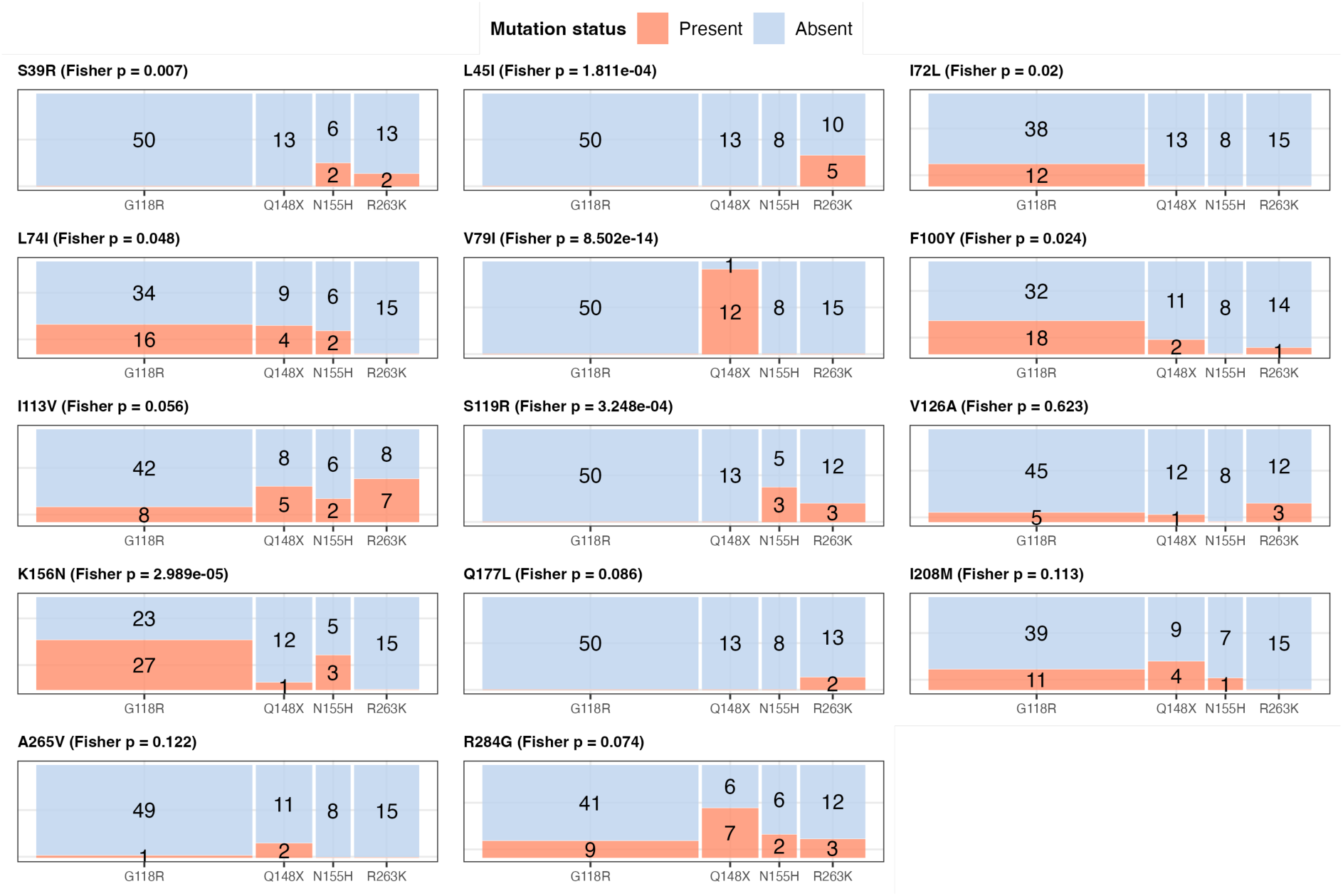
Distribution of 14 selected integrase mutations across INSTI resistance pathways. Fisher’s exact test was performed for each identified mutation across four major INSTI resistance pathways (G118R, Q148X, N155H, R263K) in sequences with DTG resistance. Q148X refers to Q148HKR pathways. Sequences with each mutually exclusive pathway were included (n = 86), while those with more than two mixed pathways were excluded. Mutation status is categorized as absent (red) and present (blue), and the size of each rectangle is proportional to the number of sequences in each category. The number of sequences in each rectangle is shown, except when it is 0. Panel titles display the mutation name and the p-value from Fisher’s exact test, indicating whether the distribution differs significantly among pathways. INSTI = integrase strand transfer inhibitor. DTG = dolutegravir.

### Comparison with Los Alamos sequences

For this analysis, we included 1,436 sequences in total (373 viraemic DTG RESIST sequences and 1,119 drug-naïve controls from Los Alamos). The comparison with the drug-naïve sequences showed that 22 other integrase mutations (S24D, V32I, K42Q, M50IR, V54I, S57G, I72L, L74I, V79I, K111Q, V126A, I135L, K156N, K160T, S195T, T128I, D232E, I251L, M275V, and S283DG) were associated with viraemia under DTG-based treatment (Supplementary Figure 5). Of these, five mutations (I72L, L74I, V79I, V126A, and K156N) had been identified as associated with genotypic DTG resistance in the previous analyses. The other 17 mutations, although associated with DTG-exposed viraemic sequences, were not enriched in those exhibiting DTG resistance.

## Discussion

In this study, we identified 14 integrase mutations that are not classified by Stanford HIVdb as major or accessory DRMs but are strongly associated with genotypic DTG resistance in this dataset of non-B subtype viruses. Most of these mutations were consistently overrepresented in resistant sequences compared to non-resistant ones. Their associations with resistance persisted after adjusting for clinical factors and were robust across HIV-1 subtypes. Notably, eight of the 14 mutations were linked to specific signature DTG resistance mutations, and five were enriched in DTG-exposed viraemic sequences, suggesting a potential role in sustaining viral replication under drug pressure.

Several of the 14 mutations have previously been proposed as compensatory. For example, the K156N mutation has been characterized in vitro as a potential accessory mutation that enhances DTG resistance when co-occurring with the major INSTI DRM N155H.^14^ In line with it, our study found that the K156N mutation is also associated with other signature major INSTI DRMs, particularly G118R, highlighting that its potential importance not limited to the N155H pathway but much broader as a compensatory mutation. Additionally, L74I has been linked to virological failure when combined with other mutations.^15^ L74I has also been recognized as a signature mutation in HIV-1 subtype A6, where it may restore replication fitness in the presence of major INSTI DRMs, particularly in the context of long-acting INSTI cabotegravir.^12^ Although the A265V mutation is known to occur naturally as a polymorphism across different HIV-1 subtypes,^11,28^ our study found it to be negatively associated with DTG resistance. Further work is required to understand whether and how its presence, in the absence of a compensatory role, might interfere with resistance development.

The majority of the identified mutations showed no significant pairwise associations, suggesting they tend to emerge independently in DTG-resistant sequences, unlike the structured pathways observed with major INSTI DRMs.^8,10^ Nonetheless, eight of the 14 mutations were linked to established INSTI resistance pathways, indicating that while they may arise independently, they could still converge on key mechanisms of DTG resistance. To date, no study has systematically examined the impact of combinations of other mutations on resistance dynamics. An analysis of the Italian ICONA cohort reported that the most frequently observed pattern among individuals experiencing virological failure involved the L74I mutation.^13^ However, only a few mutation combinations were detected, underscoring that our understanding of potential synergistic effects among these mutations in shaping resistance and viral fitness is limited.

Interestingly, we observed that while some other integrase mutations were associated with both DTG resistance and viraemia (I72L, L74I, V79I, V126A, and K156N), others were linked exclusively to one of these outcomes. Although their impact on viral fitness landscape would be limited to interpret from our findings, these distinct groups of mutations may reflect different resistance mechanisms in HIV-1, suggesting that resistance and viral replication, while interconnected, do not always overlap.

A strength of our study is the inclusion of, to the best of our knowledge, the largest dataset of non-B HIV-1 subtype sequences of PWH with virological failure on DTG-based ART, which have been underrepresented in previous studies. This is particularly relevant given prior reports of the different prevalence of other integrase mutations between subtype B and non-B viruses.^29,30^ However, our findings rely exclusively on genotypic resistance data, without accompanying phenotypic resistance, drug-level measurements, or robust adherence data. Such data would help to clarify the impact of the identified mutations on viral fitness and resistance dynamics. Other study limitations include: its cross-sectional design, sequencing limited to viral loads above 1,000 copies/mL, and the absence of pre-DTG regimen sequences which restricted our ability to capture early mutational events in DTG resistance and assess the influence of background resistance, such as nucleoside reverse transcriptase inhibitor-associated mutations, on DTG resistance. Their absence also limits our ability to assess the selective pressures driving resistance, as some mutations may have reverted over time while individuals had persistent viraemia.

In conclusion, our analysis suggests that several other integrase mutations may be associated with DTG resistance and viraemia during DTG-based treatment. Although their effects on viral fitness remain uncertain, they could represent compensatory mutations reducing the fitness cost of major resistance mutations. Phenotypic resistance testing may validate their functional roles and contributions to resistance dynamics.

## Acknowledgements

We thank all participants and healthcare providers involved in the DTG RESIST study of International Epidemiology Databases to Evaluate AIDS (IeDEA).

## DTG RESIST study group

*Argentina: Administration (Cinthia Sapienza); Clinical Staff (Camila Valeriano, Carolina Perez, Estefania Rodriguez, Florencia Cahn, Maria Victoria Iannantuono, Patricia Patterson, and Victoria Viera); Co-PI (Carina César); Data Management (Nicolás Doudtchitzky); Laboratory Staff (Ana Gun and Mariana Ferrari); PI (Pedro Cahn); Study coordination (Horacio Beylis and Lara Vladimirsky)*

*Brazil: Administration (Beatriz Leonardo, Claudia Silva, Gabriel Silva, Manoel Filho, Marcella Barboza, Sue Lima, Tania Krstic, and Ubiraçan Rufino); Co-PI (Livia Ferreira); Data Management (Alexandre Souza, Ana Carolina Figueiredo, Ana Claudia Silva, Flavia Lessa, Luiz Camacho, Ronaldo Moreira, and Thadeu Pinheiro); Laboratory Staff (Ellen Gomes, Flavia Gomes, Liriell Cordeiro, Michelli Gonçalves, Sandro Costa, and Soraia Moura); PI (Sandra Cardoso); Research (Barbara Viggiani, Daniel Arabe, and Davila Silva)*

*Burkina Faso: Administration (Sidia Arlette Sanou); Clinical Staff (Sanata Koala, Sidbéwindin Richard Ramde, and Stéphane Sanou); Data Management (Gbolo Pooda); Laboratory Staff (Abdoul-Salam Ouedraogo, Maxime Damolga, and Yacouba Sawadogo); PI (G.e. Armel Poda); Psychosocial counselor (Micheline Sanou)*

*Cambodia: Clinical Staff (Mengsomanythd Chhay, Narom Prak, Seila Pech, Sophea Heng, and Sreypov Heam); Co-PI (Mengsomanythd Chhay); Data Management (Chanthy Pov, Sopanha Pich, and Sophea Heng); Laboratory Staff (Chandara Mom and Sopanha Pich); PI (Vohith Khol)*

*Cameroon: Administration (Clarisse Lengouh); Clinical Staff (Djenabou Amadou, Eric Ngassam, Ivon Nchang, and Phyllis Fon); Data Management (Mabou Gabriel and Marc Lionel Ngamani); IeDEA Coordination (Jordanne Ching and Judith Nasah); PI (Anastase Dzudie); Study coordination (Peter Vanes Ebasone)*

*Cote d’Ivoire CNTS: Clinical Staff (Arlette Kouamé Epouse Kouadio, Bades Isidore Bohouo, Hosihiri Lambert Dohoun, and You Linda Niagne Epouse Téhé); Co-PI (Kouadio Stéphane N’goran); Data Management (Konan Raoul Kouakou); Laboratory Staff (Mathias Balie and Vincent Camille Sablin); PI (Kla Albert Minga)*

*Cote d’Ivoire CePREF: Clinical Staff (Amah Cécile Tchehy and Jeannot Goli); Co-PI (Anzian Amani); Data Management (Emma Nadège Kokogny and Issouf Koffi Ladji); Laboratory Staff (Sammuel Assande); PI (Eugène Messou); Research (Karidiatou Diallo)*

*Durban Lab: Co-PI (Jennifer Giandhari and Tulio De Oliveira); Data Management (Bertha Baye and Hastings Musopole); Laboratory Staff (Lavanya Singh, Nonkululeko Avril Mbatha, Samukelisiwe Pretty Khathi, Shirelle Naidoo, Sureshnee Pillay, and Xolani Hilorious Zulu); PI (Richard John Lessells)*

*India: Administration (Beulah Balakrishnan); Clinical Staff (Dr.keerthana Priya Thamayanthi Shankar, Esther Mony, and Ramya Murugan); Co-PI (Dr.poongulali Selvamuthu); Data Management (Blessy Bobby, Jacinth Sugumar, Rajesh Elangovan, Sasi Jothiramalingam, and Shilpa Jose); Laboratory Staff (Dr. Priya Kannian, Gracemary Arul, Harika Paila, Mahanathi Pasuvaraj, Shalini Loganathan, and Sivaranajani Durairaj); PI (Dr. Kumarasamy Nagalingeswaran)*

*Kenya: Clinical Staff (Cosmas Apaka and Julius Cheruiyot); Co-PI (Shamim M. Ali and Suzanne Goodrich); PI (Kara Wools-Kaloustian and Lameck Diero)*

*Malawi: Administration (Joseph Chintedza); Clinical Staff (Erick Mtemang’ombe, Jessie Hau, Kelvin Rambiki, and William Maliko); Co-PI (Jacqueline Huwa, Rose Nyirenda, and Wilson Bilaal); Data Management (Blessings Mwandira, Geldert Chiwaya, and Pachawo Bisani); Laboratory Staff (Enock Khunju, Rafiq Maluwa, and Shameem Buleya)*

*Mexico Lab: Data Management (Claudia Garcia-Morales); Laboratory Staff (Margarita Matias-Florentino); PI (Santiago Avila-Rios) Rwanda: Administration (Marie Gertrude Bahire Rutwaza); Clinical Staff (Fabienne Shumbusho, Francine Umwiza, Samuel Munyentwari, and Verene Mukankurunziza); Co-PI (Marcel Yotebieng); Data Management (Benjamin Muhoza); Laboratory Staff (Faustin Kanyabwisha); PI (Gad Murenzi)*

*Switzerland: Data Management (Mamatha Sauermann, ipek Çelikağ, Tom Loosli, Nuri Han, and Gioia Wick); PI (Matthias Egger and Roger D Kouyos)*

*Thailand HIVNAT: Co-PI (Napon Hiranburana); Data Management (Chuleeporn Wongvoranet and Penpanat Toomcharoen); Laboratory Staff (Sasiwimol Ubolyam); PI (Anchalee Avihingsanon)*

*Thailand Lab: Laboratory Staff (Suwanna Mekprasan); PI (Sunee Sirivichayakul) Thailand Ramathibodi: Clinical Staff (Laor Nakgul and Nutaporn Sanmeema); Co-PI (Asoc.angsana Phuphuakrat); Data Management (Laor Nakgul and Nutaporn Sanmeema); Laboratory Staff (Laor Nakgul and Nutaporn Sanmeema); PI (Prof.sasisopin Kiertiburanakul)*

*Uganda Masaka: Clinical Staff (Aloysius Ssabayinda, Dennis Madanda, Monica Nakirya, Samuel Katungi, Stella Nabunnya, and Wilson Kazoora); Data Management (Matthew Ssemakadde and Phoebe Mutenyo); Laboratory Staff (Moses Asiimwe); PI (Charles Kasozi and Lydia Buzaalirwa)*

*Uganda Mbarara: Administration (Bronia Mwiine); Clinical Staff (Alexis Byaruhanga, Bob Ssekyanzi, Caroline Kusingura, Lillian Ayesiga, and Sarah Namwanje); Co-PI (Helen Byakwaga); Data Management (David Muhumuza); Laboratory Staff (Yona Mbalibulha); PI (Winnie Muyindike)*

*Zambia: Clinical Staff (Alice Miyanda, Aretha Mumba, Fiona Mureithi, Kenan Simumba, and Vivian Tonga); Co-PI (Guy K. Muula); Data Management (Caroline Chileshe, Esau Banda, Ethel Muyanga, Jackson Daka, Josephine Mboozi, Suwilanji Nalungwe, and Sydney Kamiji); Laboratory Staff (Chenge Mukonde, Choolwe Bwalya, Paul Pandala, and Tabiso Mubiana); PI (Carolyn Bolton); Peer educator (Pamela Kabombo and Thandiwe Phiri)*

*Zimbabwe: Clinical Staff (Paddington Marume, Varaidzo Kachingwe, and Wilson Marikopo); Data Management (Ardele Mandiriri, Maureen Wellington, Sydney Malunga, and Varaidzo Moyo); Laboratory Staff (Dakarayi Magumise and Ratidzai Katsidzira); PI (Cleophas Chimbetete)*

## Data availability

The sequence data are available in GenBank under accession numbers PV008448-PV008617, PV419540-PV419544, PV419550, PV419551, PV419554, PV419555, PV419557-PV419583, PV467483-PV467495, PV816612-PV816623, PV816625, PV816629-PV816633, PV816635-PV816650, PV816652-PV816682, PV816684, PV816687, PV816689-PV816692.

## Transparency declarations

H. F. G. has received grants from the Swiss National Science Foundation, the Swiss HIV Cohort Study, the Yvonne Jacob Foundation, University of Zurich’s Clinical Research Priority Program, Zurich Primary HIV Infection, Systems. X, the Bill C Melinda Gates Foundation, the US NIH, Gilead Sciences, ViiV Healthcare, and Roche; has received honoraria for advisory boards from Gilead Sciences, ViiV Healthcare, Janssen, GSK, Johnson C Johnson, and Novartis; has received honoraria for a data safety monitoring board from Merck; and has received a travel grant from Gilead Sciences. R. L. has received financial support (payment to the institution) for the present work from the NIH NIAID and has received grants (payment to institution) from the NIH NIAID for an unrelated project. R. D. K. receives grants from the Swiss National Science Foundation, the US NIH, and Gilead Sciences. All other authors: none to declare.

