## Supplementary Material for "Novel integrase mutations linked to genotypic DTG resistance in non-B HIV-1 strains from African participants: The DTG RESIST study"

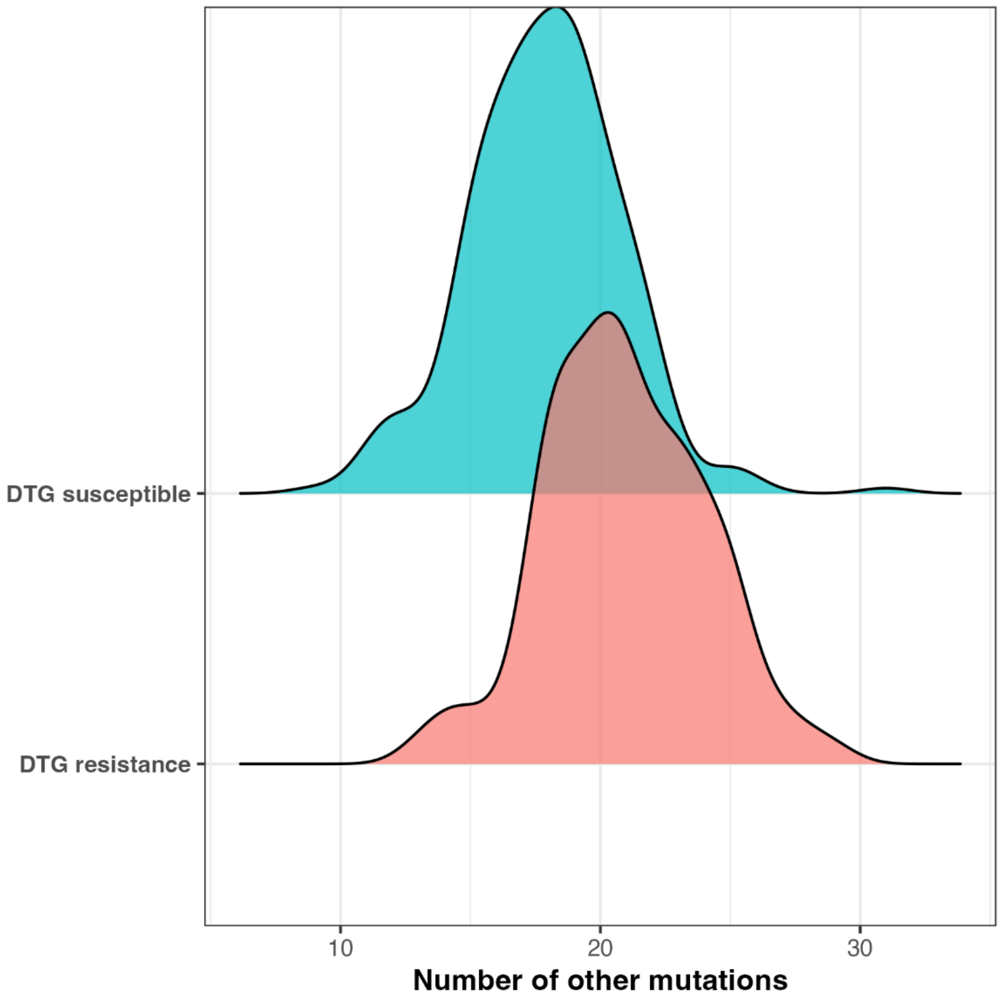


**Supplementary figure 1 Density of number of other integrase mutations from sequences with DTG resistance (red) and sequences without DTG resistance (blue).** Welch two sample t-test was performed to compare the mean number of other mutations between sequences with DTG resistance ("DTG resistance") and those without ("DTG susceptible"): t-statistics = -8.86 [95% confidence intervals: -3.73 to -2.37], p-value = 6.978e-16. DTG = dolutegravir.

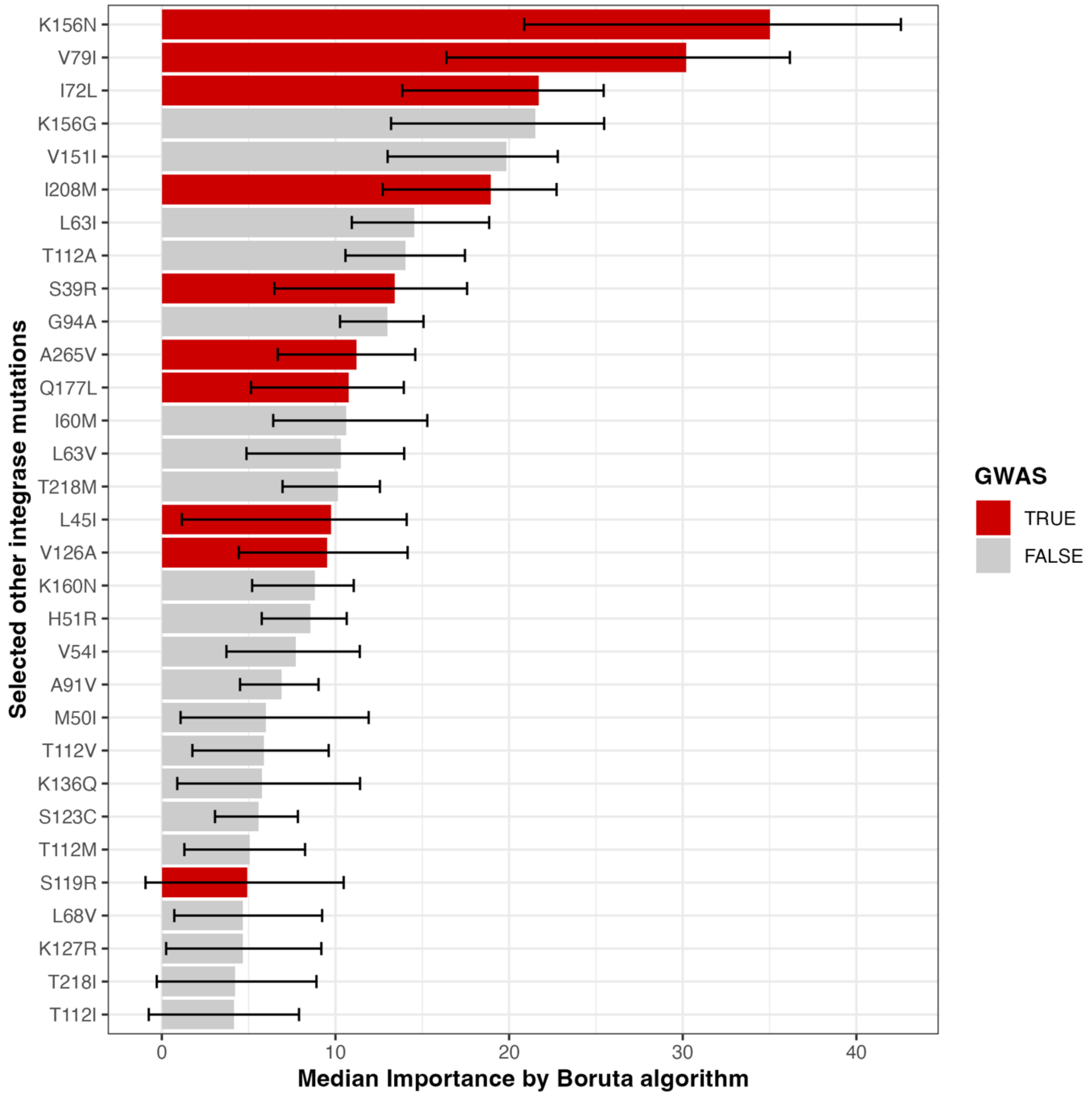


**Supplementary Figure 2 Median importance scores for integrase mutations calculated using the Boruta algorithm.** The Boruta algorithm identifies features relevant for predicting DTG resistance. Mutations classified as confirmed by the Boruta algorithms are listed, representing mutations which importance scores were significantly above permuted mutations. Higher median importance scores indicate greater predictive relevance. If they have been identified by GWAS to be associated with DTG resistance, they are coloured by red. Mutations previously identified by GWAS are coloured by red. DTG = dolutegravir.

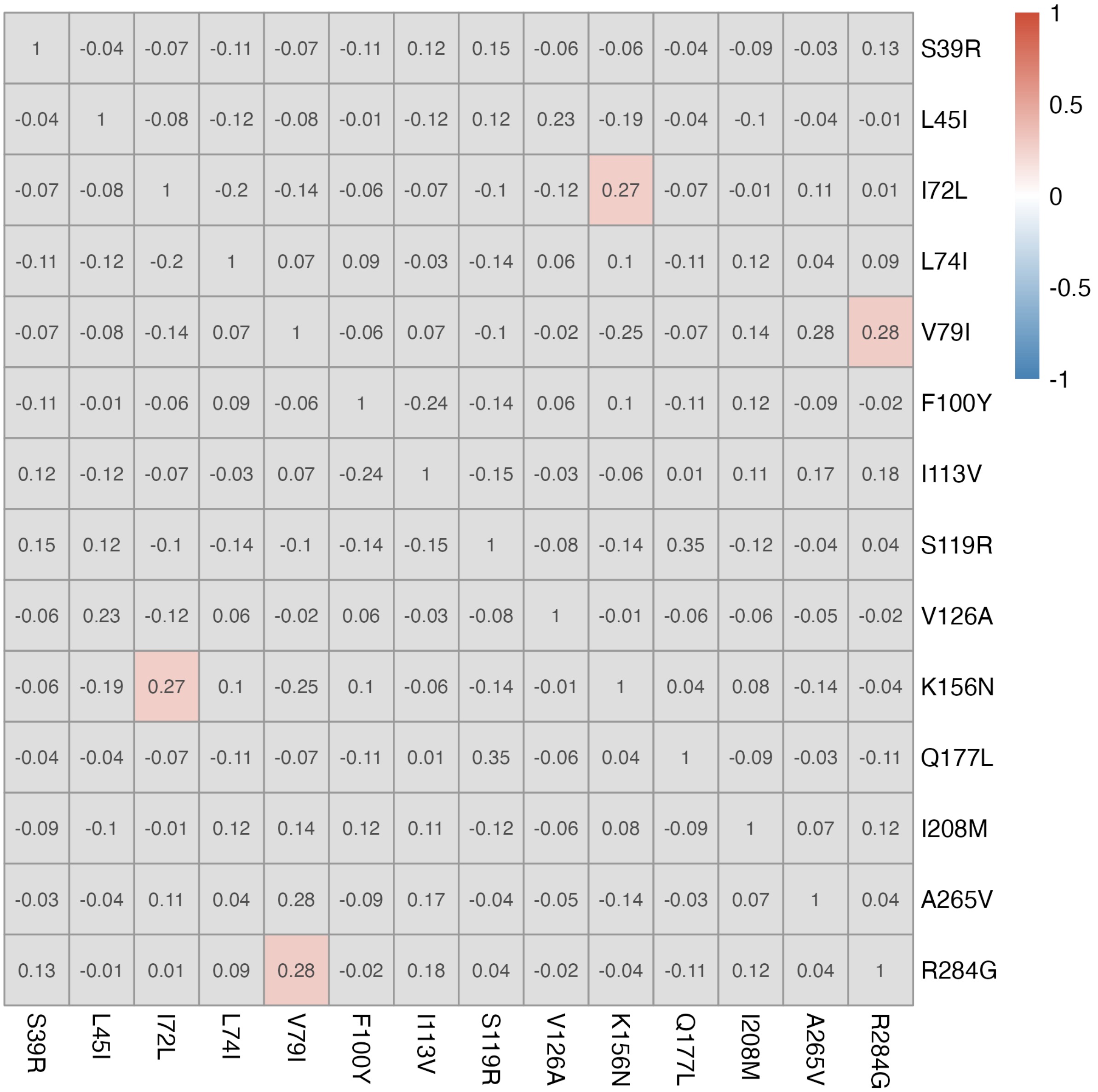


**Supplementary Figure 3 Pairwise Phi coefficients among selected mutations in sequences with DTG resistance.** The Phi coefficient (ϕ) quantifies the strength and direction of linear relationships between selected other integrase mutations. Mutations identified from GWAS firth regression were included in the analysis. Non-significant associations (BH-adjusted p-value > 0.05) were marked as grey. P-values were derived from contingency table tests: Fisher's exact test was used when cell counts were < 5, and Pearson's chi-squared test was applied otherwise. Red indicates strong positive co-occurrence, while blue indicates negative co-occurrence (i.e., their occurrence is independent). DTG = dolutegravir. GWAS = genome-wide association study. BH = Benjamini-Hochberg.

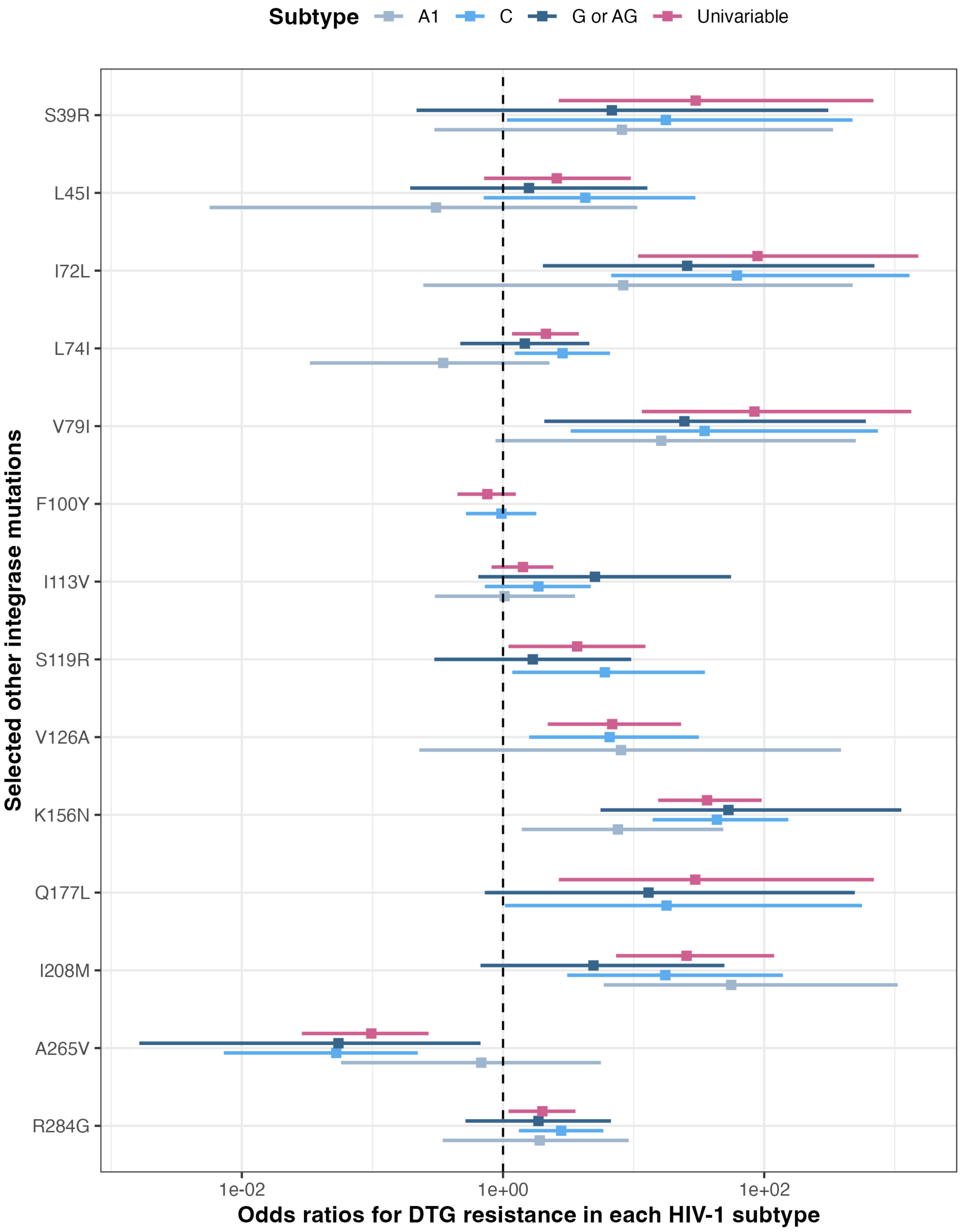


**Supplementary Figure 4 Odds ratios for DTG resistance in HIV-1 subtypes.** Odds ratios with 95% credible intervals were calculated using a Bayesian regression model. Mutations identified from GWAS firth regression were tested within each subtype. Among them, mutations absent in each subtype were excluded in the analysis with the respective subtype. Each model in each subtype is shown in different colours. DTG = dolutegravir. GWAS = genome-wide association study.

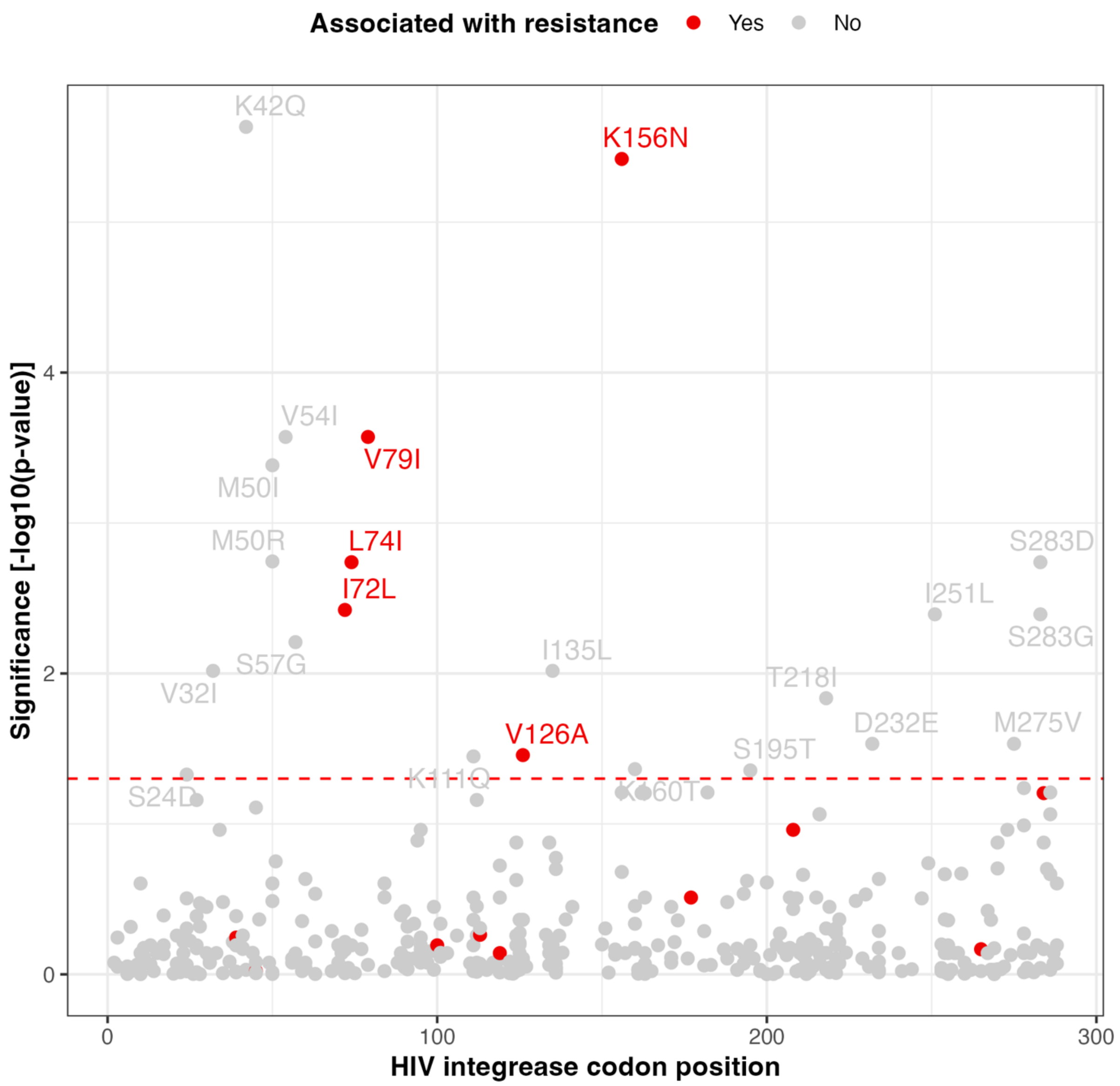


**Supplementary Figure 5 Genome-wide association study (GWAS) of other integrase mutations and their associations with viraemic sequences under DTG-based treatment.** A case-control design (1:3 matching) was used to select sequences with the lowest genetic distance for the analysis. For GWAS, Firth logistic regression was applied, including the first 10 PCs and subtypes to account for population structure. P-values were adjusted using the BH procedure, with a significance threshold of adjusted p-value < 0.05. The name of significant mutations were displayed and mutations identified in the

GWAS for association with DTG resistance (Figure 1) are highlighted in red. DTG = dolutegravir. PC = principal component. BH = Benjamini-Hochberg.

**Supplementary Table 1. Other integrase mutations associated with DTG resistance.** 25 other integrase mutations are listed, each significantly overrepresented in sequences with DTG resistance, using fisher's exact test ('Odds ratios [95% CI]' and 'P-value'). P-value was adjusted for multiple comparisons using the Benjamini-Hochberg procedure. Number of each mutation in sequences with and without DTG resistance is described in the first two columns. DTG resistance, here, is defined as having at least one intermediate level of major DTG DRMs. DTG = dolutegravir. DRMs = drug resistance mutations.

| Mutation | Number in sequences without DTG resistance | Number in sequences with DTG resistance | Odds ratios [95% CI] | P-value | GWAS |
|---|---|---|---|---|---|
| *V151I* | 1 | 17 | 52.17 [7.96, 2184.16] | <0.001 | No |
| *K156N* | 5 | 44 | 37.97 [14.29, 127.96] | <0.001 | Yes |
| *I208M* | 2 | 18 | 27.85 [6.47, 250.69] | <0.001 | Yes |

| | | | | | |
|---|---|---|---|---|---|
| *L63I* | 5 | 22 | 14.12 [5.02, 49.15] | <0.001 | No |
| *T112A* | 5 | 21 | 13.32 [4.7, 46.58] | <0.001 | No |
| *T112M* | 2 | 7 | 9.66 [1.8, 96.87] | 0.022 | No |
| *L63V* | 3 | 8 | 7.41 [1.74, 44.27] | 0.021 | No |
| *V126A* | 4 | 10 | 7.07 [1.98, 31.62] | 0.007 | Yes |
| *V54I* | 4 | 9 | 6.3 [1.71, 28.66] | 0.019 | No |
| *I60M* | 10 | 17 | 5.07 [2.11, 12.88] | 0.001 | No |
| *M50I* | 71 | 43 | 1.97 [1.2, 3.25] | 0.043 | No |
| *T218I* | 105 | 20 | 0.38 [0.21, 0.67] | 0.005 | No |

| | | | | | |
|---|---|---|---|---|---|
| *T112V* | 238 | 71 | 0.35 [0.2, 0.61] | 0.002 | No |
| *A265V* | 65 | 3 | 0.1 [0.02, 0.3] | <0.001 | Yes |
| *I72L* | 0 | 13 | Inf [8.8, Inf] | <0.001 | Yes |
| *V79I* | 0 | 13 | Inf [8.8, Inf] | <0.001 | Yes |
| *S123C* | 0 | 4 | Inf [1.76, Inf] | 0.042 | No |
| *T218M* | 0 | 6 | Inf [3.18, Inf] | 0.005 | No |
| *K160N* | 0 | 4 | Inf [1.76, Inf] | 0.042 | No |
| *G94A* | 0 | 11 | Inf [7.11, Inf] | <0.001 | No |
| *K156G* | 0 | 11 | Inf [7.11, Inf] | <0.001 | No |
| *A91V* | 0 | 4 | Inf [1.76, Inf] | 0.042 | No |

| | | | | | |
|---|---|---|---|---|---|
| *Q177L* | 0 | 4 | Inf [1.76, Inf] | 0.042 | Yes |
| *H51R* | 0 | 6 | Inf [3.18, Inf] | 0.005 | No |
| *S39R* | 0 | 4 | Inf [1.76, Inf] | 0.042 | Yes |
